# Foundation time series models for forecasting and policy evaluation in infectious disease epidemics

**DOI:** 10.1101/2025.02.24.25322795

**Authors:** Suprabhath Kalahasti, Benjamin Faucher, Boxuan Wang, Claudio Ascione, Ricardo Carbajal, Maxime Enault, Christophe Vincent Cassis, Titouan Launay, Caroline Guerrisi, Pierre-Yves Boëlle, Federico Baldo, Eugenio Valdano

## Abstract

Epidemic forecasting and policy evaluation rely on mathematical models to predict infectious disease trends and assess the impact of public health policies. Traditional models typically require extensive epidemiological data and may struggle in data-limited settings. Transformer-based, foundation AI models have demonstrated strong predictive capabilities in various time series applications. We investigated whether they can be the basis of a new epidemic modeling framework. We evaluated five foundation models - TabPFN-TS, TimeGPT, TimesFM, Lag-Llama, and Chronos - across diverse pathogens, diseases and locations, including influenza-like illness, RSV, chickenpox, dengue, COVID-19 and neonatal bronchiolitis. Models were tested for long-term forecasting (multi-season predictions), short-term forecasting (four-week-ahead predictions), and epidemic peak timing estimation. We also assessed their ability to generate counterfactual scenarios in policy evaluation, using COVID-19 restriction measures in Italy, RSV immunization in France, and synthetic epidemic data as validation. Foundation models demonstrated strong predictive accuracy, possibly outperforming traditional statistical and mechanistic models in data-limited contexts. They generated multi-season forecasts and short-term forecasts with good accuracy and stable uncertainty. They gave reliable peak timing estimates months before the actual peak. In policy evaluation, TabPFN-TS accurately estimated intervention effects, matching estimates from an independent epidemiological study. Our findings suggest that foundation models can complement existing approaches in epidemic modeling. Their ability to generate accurate forecasts and counterfactual analyses with minimal data input highlights their potential for real-time public health decision-making, particularly in emergent and resource-constrained settings. Further research should explore domain-specific adaptations to optimize performance for infectious disease modeling.

## Introduction

Mathematical models provide essential support to public health response to epidemics of communicable diseases. They forecast epidemic indicators such as incident cases and hospitalizations to assess epidemic trends and healthcare system capacity, while scenario-based analyses help identify the most effective intervention strategies.

Epidemic models have found applications across diseases, transmission routes and locations. Major public health agencies routinely coordinate forecasting efforts for seasonal acute respiratory illness^1,2^. Similar efforts extend to vector-borne endemic diseases, such as dengue^3^ and West Nile Virus^4^. Models were used to predict local transmission and global diffusion of potential and actual pandemics, including the 2009 H1N1 influenza pandemic^5^, the 2014-2016 West Africa Ebola outbreak^6^, the 2016-2017 Zika pandemic^7^, and most recently, the COVID-19 pandemic^8,9^. Mathematical models also routinely help select among alternative policies by estimating their projected effectiveness and costs, from HIV/AIDS^10^ to malaria^11^. During COVID-19, models helped evaluate and design nonpharmaceutical interventions^12^ and vaccination campaigns^13^.

Despite their active development and widespread use, epidemic models face critical limitations in their predictive power and applicability. As public health needs require increasing accuracy and predictive power, models have relied more and more heavily on large-scale data tracking epidemic indicators and drivers, such as nontraditional surveillance^14^, genomic data^15^, host contact patterns and mobility^8,16^, climate and the environment^3^. But these data-intensive models struggle to generalize beyond the specific epidemiological context under study, as they are often tailored to multiple ad-hoc fit for purpose data feeds that may not be available everywhere, at all times^17,18^. Their performance is thus constrained by context-specific and epidemic-specific data availability, which is heterogeneous and cannot increase indefinitely. As opportunity determines data availability more than public health needs, the communities with the proper technological infrastructures for safe and effective data collection may reap the benefits of epidemic modeling, while, at the same time, those with loose protections of individual rights may see increased risk of sensitive data misuse^19^.

To overcome this limitation, new modeling frameworks should be able to leverage data from outside the specific epidemiological context under study and make reliable predictions with the available information. Artificial intelligence (AI) is emerging as a potential methodological approach to achieve this goal^20^. In this context, most current efforts combine traditional mechanistic models with deep learning^21^. This is based on the idea that constraining deep learning models with domain-specific knowledge in the form of the elementary processes of disease spread (e.g., transmission, severity, immunity) can combine the explainability and identifiability of mechanistic models with the ability of deep-learning to extract complex information patterns better than traditional statistical inference, to enhance predictive performance^22^.

Here, we instead adopted a different approach to integrating AI into epidemic modeling, based on the recent evidence that foundation AI models based on transformer architectures perform very well on tabular and time series data^23^. Foundation models^24^ are based on deep neural networks and self-supervised learning. They are trained on broad datasets, with the scope to fine tune them and use them for downstream tasks. Foundation models in the form of Large Language Models (LLM) have revolutionized natural language processing, de facto opening the era of consumer-available Artificial Intelligence. These models excel in transferring knowledge across domains, leveraging patterns learned from datasets from completely different domains to enhance forecasting accuracy and content generation even in data-scarce settings^25,26^.

In this study, we investigated whether existing foundation models for time series prediction can form the basis of a new architectural framework for epidemic modeling. We tested their performance across different epidemics and diseases, and on different predictive windows and tasks. We tested four models that extend major LLM architectures - Google’s TimesFM^27^, Meta’s Lag-Llama (based on LLaMA)^28,29^, Amazon’s Chronos (based on T5 architecture)^30,31^, TimeGPT (based on self attention mechanism transformers)^32,33^ - and TabPFN^34–36^, a model built for tabular data. Notably, none of the models we used was ever designed or trained for epidemic modeling, nor for use in public health and epidemiology. Notwithstanding, we tested their performance on epidemic time series without any major retraining, fully leveraging the dramatic power of generalization across knowledge domains of transformer-based models. We compared their performance with established epidemic and statistical models, demonstrating their potential to improve the reliability of epidemic forecasting and policy evaluation, particularly in data-limited settings, where they may outperform traditional approaches. Additionally, their ability to function with minimal fine-tuning - what is known as few-shot or zero-shot learning - significantly reduces computational demands: These models can run efficiently on standard hardware, such as a laptop, making them well-suited for both resource-constrained and data-scarce environments.

## Methods

### Data

Surveillance data of Influenza-like illness (ILI) in France from 1985 to 2019 were collected through the country’s sentinel surveillance system *Réseau Sentinelles*^37,38^. This system uses a network of general practitioners distributed across the country who report the number of ILI cases observed in consultations on a weekly basis. ILI cases are defined as a sudden onset of fever exceeding 39°C, with myalgia or respiratory symptoms. Each week, a subset of these cases undergo virological testing for various respiratory viruses. Test results were then used in combination with ILI data to estimate the incidence of influenza and respiratory syncytial virus (RSV) since 2013 as done in Ref^39^. Chickenpox incidence data were also obtained from the same system. A chickenpox case was defined as presenting the characteristic rash (erythematous-vesicular eruption lasting 3-to-4 days, pruritic, followed by a drying phase) with a sudden onset of mild fever (between 37.5 and 38°C). Dengue incidence data in Brazil came from Ref.^37^.

COVID-19 incidence data by region in Italy, from November 2020 to May 2021, came from Ref.^40^. During that period, Italy implemented a tiered system of restrictions, classifying regions into white (rarely witnessed) yellow, orange, and red, based on epidemiological risk^41^. Yellow tier enforced light restrictions, with limited curfews and indoor dining allowed, while orange imposed stricter measures, including travel restrictions between municipalities and the closure of bars and restaurants. The red tier had the most severe restrictions, including stay-at-home orders, school closures, and the shutdown of non-essential businesses. The tier classification was determined automatically based on a set of epidemiological indicators, including the weekly incidence rate per 100,000 inhabitants, ICU occupancy rates, and the reproduction number (Rt), with stricter measures triggered as thresholds were exceeded^41^.

Bronchiolitis admissions to the ER in children less than 1 year-old came from seven pediatric hospitals in the region of Paris, France, over 5 seasons (October-February; years 2017-2019, 2022-2023). The last season saw the wide scale introduction of nirsevimab, a monoclonal antibody providing passive immunization against bronchiolitis caused by RSV. Immunization was rolled out in September 2023 among children who were less than 6 months-old at this date, then at birth from October 2023 to December 2023. Coverage was assessed among children visiting the ER for conditions other than bronchiolitis^42^. Figure 1 shows all the data available to this study and Tab. 1 summarizes their characteristics.

**Figure 1:**
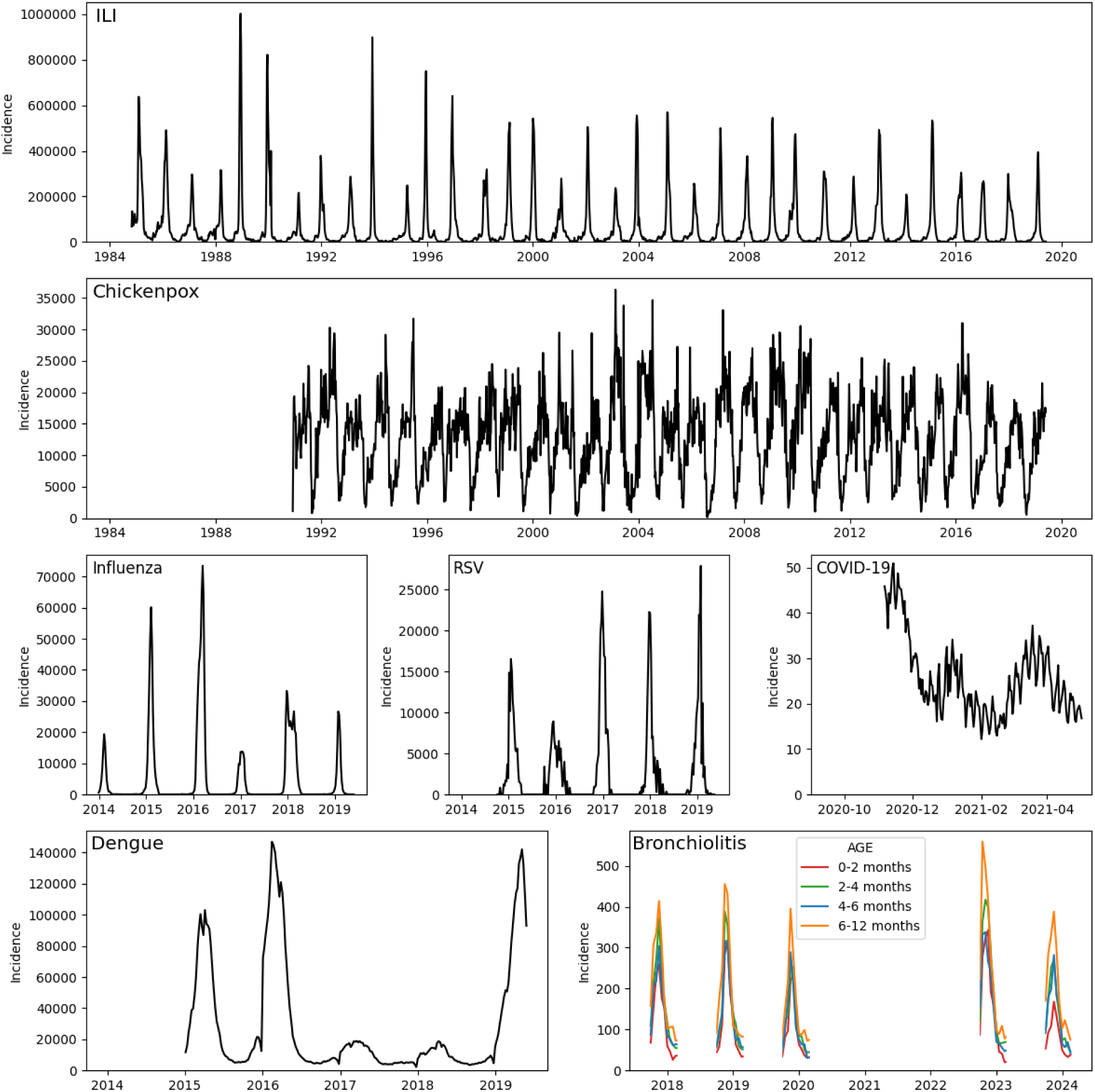
Datasets used in this study. For COVID-19, only the region of Latium is represented here, see Appendix for the other. Incident cases are reported weekly for ILI, chickenpox, influenza, RSV and dengue, daily for COVID-19. Bronchiolitis-related ER admissions are reported fortnightly.

**Table 1:** Data description. Summary features of the different datasets used.

| Dataset | 1st data point | Last data point | # data points | # seasons | Median | Q1 | Q3 | Max | Loc | Freq |
| --- | --- | --- | --- | --- | --- | --- | --- | --- | --- | --- |
| ILI | 1984-10-29 | 2025-01-20 | 2092 | 41 | 17426 | 5449 | 53574 | 1001824 | France | weekly |
| Influenza | 2013-12-30 | 2024-01-29 | 527 | 6 | 30 | 0 | 872 | 73442 | France | weekly |
| RSV | 2014-10-06 | 2024-01-29 | 370 | 5 | 373 | 0 | 2509 | 27912 | France | weekly |
| Chickenpox | 1990-12-03 | 2025-01-20 | 1771 | 35 | 12034 | 6704 | 16670 | 36298 | France | weekly |
| Dengue | 2015-01-04 | 2022-12-25 | 413 | 7 | 13716 | 6801 | 33903 | 151784 | Brazil | weekly |
| COVID-19 | 2020-11-06 | 2021-05-02 | 178 | 1 | 23 | 19 | 30 | 51 | Italy | daily |
| Bronchiolitis | 2017-10-01 | 2024-02-16 | 200 | 5 | 112 | 69 | 221 | 559 | Paris, France | fortnightly |

### Benchmark models

We used three established models as benchmark: two statistical models and one mechanistic model. The first statistical model is Seasonal Autoregressive Integrated Moving Average (SARIMA)^43^ as implemented in the R package in Ref.^44^. For each scenario, the model was trained on the available data up to the last point of the training data set. The hyperparameters were calibrated using the auto.arima function. The second statistical model is the time series analysis model Prophet developed by Meta^45^. The mechanistic model is similar to what was used in Ref.^39,46,47^, fitted using Markov Chain Monte Carlo. More details are given in the Appendix.

### Foundation models

#### TabPFN-TS

An extension of the Tabular Prior-data Fitted Network (TabPFN^34–36^) - a transformer-based bayesian inference model - TabPFN-TS frames a time series forecasting problem as a regression problem within tabular data. The model uses features extracted from the timestamps within the dataset such as sine and cosine encodings and calendar indices to capture widespread time scales.

**<u>Chronos</u>** - specifically Chronos Bolt (Small) - is a probabilistic time series forecasting model^30^. It converts time series value into tokens using scaling and quantization techniques. The tokens are converted into a corpus and are then trained on existing transformer based language model architectures through cross-entropy loss. Chronos models are based on T5 (Text-to-Text Transfer Transformer)^31^ encoder-decoder architectures which use transfer learning on Natural Language Processing (NLP). Chronos was trained on a large collection of publicly available datasets in combination with synthetic datasets that were generated from Gaussian processes.

**<u>Lag-Llama</u>**^29^ from Meta is pre-trained on 27 time series datasets (7,965 univariate time series) across multiple disciplines. It consists of parameters ranging from 7 to 65 billions. The model uses an inference function that returns predictions for a given horizon based on the prediction length and the number of samples it needs to sample from the predicted probability distribution to generate forecasts. We used 100 as the number of samples the model would need to sample from for our analysis. We fine tuned its context length setting it to 64.

**<u>TimesFM</u>** is a foundation model by Google trained on a large corpus of time series gathered through Google trends/search and Wikipedia. It has 200M parameters and a pre-training size of 200B time points. From an architectural standpoint, the model uses patch based modelling^48^ in breaking down the time-series datasets during the training phase and is trained in a decoder^49^ only format resulting in patches based prediction based on previous patches. The model was configured with a per-core batch size of 32, with 50 layers, a context length of 2048, and a prediction horizon of 200 time steps. Positional embeddings were disabled to optimize performance. The model was initialized from the pre-trained checkpoint. The frequency input was set to 1, aligning with medium-frequency time series data (e.g., weekly and monthly granularity).

**<u>TimeGPT</u>** from Nixtla is a transformer-based foundation model built on self-attention mechanisms^33^ consisting of an encoder-decoder structure in place that uses past historical data to generate forecasts.The model is trained on 100 billion data points gathered from multiple publicly available datasets across different domains coupled along with information on various temporal patterns to account for seasonality in varying degrees. Conformal prediction^50,51^ frameworks were used to generate forecasts.

We stress that all models are pre-trained, and applying them to our datasets required only minimal fine-tuning, which could be done very efficiently. Specifically, fine-tuning of TabPFN, which we used for policy evaluation, could be fully performed on a laptop requiring less than one minute for instance.

### Evaluation of forecasting performance

We evaluated models on five datasets (ILI in France, influenza in France, RSV in France, chickenpox in France, dengue in Brazil) and across three forecasting tasks: long-term multi-season incidence forecasting, short-term incidence forecasting and peak timing forecasting. Forecasting scenarios spanned three seasons, from 2016 to 2019. For each scenario, all available data before the start of the forecasting window were used for model fitting or fine-tuning. This resulted in different epidemics having very different sizes of the training data set (see Fig. 1). For each scenario and model, we forecast the median and the 1st and 9th deciles, except for TimeGPT which provided point predictions only.

Long-term, multi-season forecasting consisted in stopping training at the start of the 2016-2017 season (early October for ILI, influenza, RSV and early September for chickenpox and dengue) and then predicting three seasons, up to 2019. Short-term forecasting consisted in predicting incidence four weeks into the future. For each dataset and each of the three seasons, forecast scenarios started at different time points from four weeks before the actual peak, to four weeks after. Peak timing forecasting consisted in predicting the week of the season at which case incidence would peak. We tested short-term peak timing forecasting, whereby the peak timing had to be predicted from data up to four weeks prior to the actual peak, and early peak timing forecasting, whereby peak timing was predicted from the start of the corresponding season, defined as before. For the various scenarios we also tested alternative versions whereby either the start of the season was changed (different month) or the short-term forecasting window was. All forecasting scenarios are listed in Appendix.

We evaluated incidence forecasts using Mean Absolute Error (MAE) divided by the dataset mean value (see Tab. 1) and Mean Absolute Percentage Error (MAPE), and peak timing forecasts using Absolute Error (AE). We then defined improvement in MAE (or MAPE) as the relative drop in a model’s error with respect to the mechanistic model. We also tested Prophet as reference in the Appendix. Scores are further detailed in Appendix.

### Policy evaluation

For COVID-19, we focused on the Latium region in Italy, where increasing SARS-CoV-2 incidence in February-March 2021 led to a delayed tightening of restriction measures on March 14. Using data up to February 22, 2021, we fine-tuned TabPFN-TS and estimated the impact of an earlier adoption of stricter restrictions (orange tier) by comparing the predicted incidence under this scenario with the observed trajectory, using past data from Italian regions and including the tier (yellow, orange, red) as additional covariate. The difference between observed and counterfactual incidence provided an estimate of the number of cases that could have been averted had stricter measures been implemented earlier.

For RSV immunization, we evaluated the effect of nirsevimab introduction on bronchiolitis-related emergency room (ER) admissions among infants aged 0-12 months, stratified in three age classes: 0-3 months, 4-6 months, 7-12 months. We fine-tuned TabPFN-TS on four historical pre-immunization bronchiolitis data between 2017 and 2023 and generated age-stratified forecasts for the 2023-2024 season under a counterfactual scenario in which immunization had not been introduced. We measured averted ER admissions as a difference between the model prediction and the data, and estimated the age-stratified effectiveness in reducing all-cause bronchiolitis ER admission as the relative reduction in observed admissions relative to model-predicted admissions, divided by the expected nirsevimab coverage among infants in that age class.

To validate the ability of foundation models to generate accurate counterfactuals, we also used synthetic epidemic data generated from a standard stochastic, discrete-time SEIRS model incorporating seasonality and interventions. The model tracked four compartments - susceptible, exposed, infected, and recovered individuals - under a constant population size, with transitions among compartments modeled through binomial sampling. We fine-tuned TabPFN-TS using data from synthetic regions and tested its ability to estimate the effects of both implemented and hypothetical interventions. The accuracy of these estimates was evaluated by comparing predicted and actual incidence under predefined counterfactual scenarios. More details are present in Appendix.

## Results

### Long- and short-term incidence forecasting

First, we tested the performance of foundation models to forecast incidence several seasons into the future, from October 1, 2016 to August 31, 2019, except for the mechanistic model and Chronos, which could forecast only one season. For each disease, we fitted or fine-tuned each model with all the available data prior to the start of the forecast window. Figure 2 reports the forecasts of all models and diseases. Benchmark models had a poor performance on long-term forecasts. Among them, Prophet was fairly accurate on the time series which had the longest training data - ILI and Chickenpox (see Fig. S1). Its forecast, however, had a very wide uncertainty. Lag-Llama was not able to perform long-term forecasts, Chronos only on chickenpox, with the limitation of being restricted to only one season. The picture was completely different for TimesFN, TimeGPT and TabPFN-TS. They showed remarkable performance on ILI, influenza and chickenpox where forecasts were both accurate and precise (small confidence intervals) up to three seasons into the future, despite a small bias in the timing of the peak for ILI and influenza. For RSV and dengue, TimesFM and TabPFN-TS could forecast the timing and shape of the season, but not the peak incidence. TimeGPT could do it for dengue, not for RSV.

**Figure 2:**
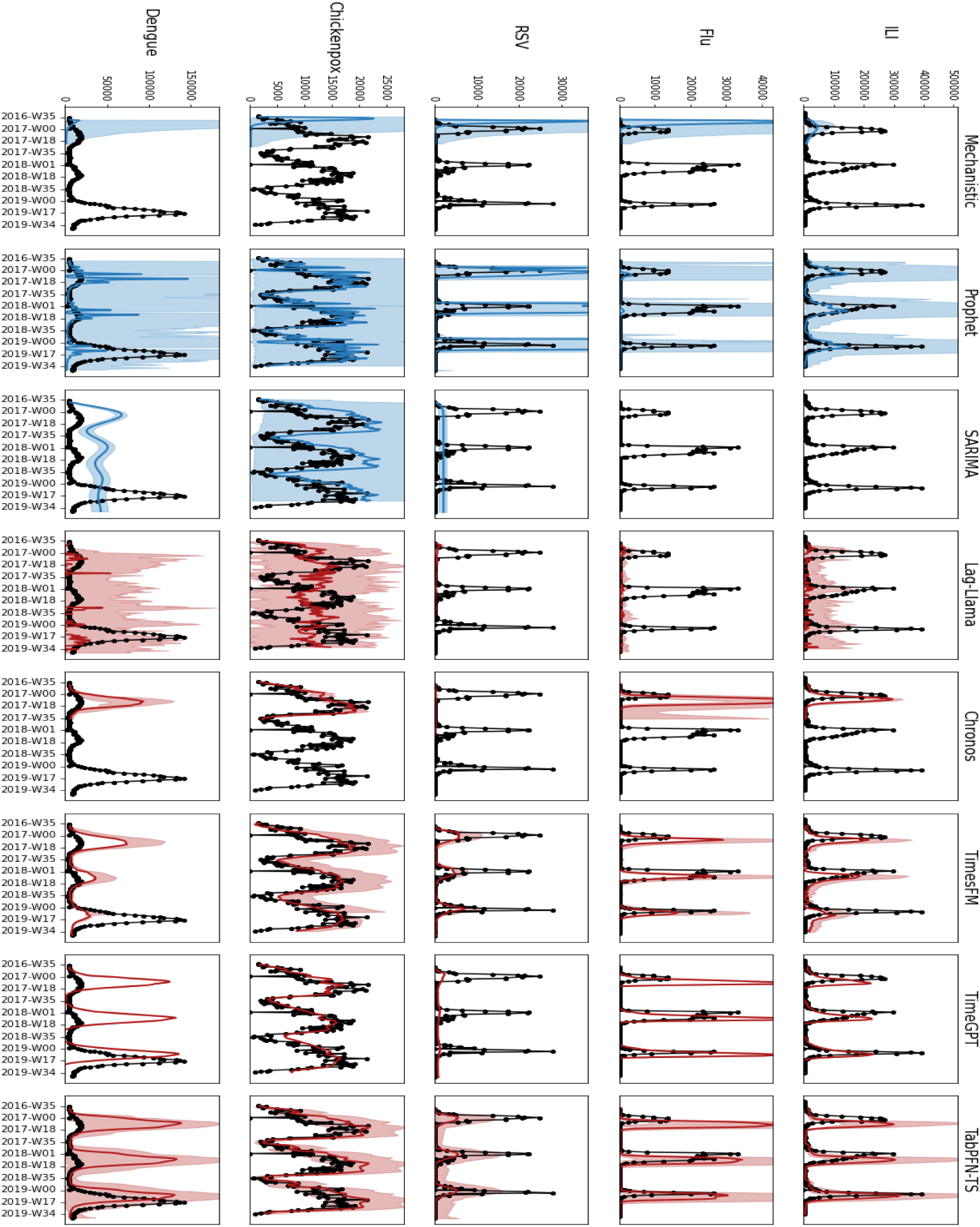
Long-term forecasting. Each plot compares the three epidemic seasons between 2016 and 2019 with model forecasts. Each row is a different dataset (disease) and each column is a different model. Benchmarks are in blue, foundation models are in red. The solid line is the median prediction, the shaded area encompasses the range between the 1st and the 9th decile. Forecasts start in October for ILI, Influenza, RSV and dengue and in September for Chickenpox.

We then tested the performance on a shorter-term forecast, namely predicting incidence four weeks into the future. Each model produced nine predictions for each disease and in each of the three seasons from 2016 to 2019. Figure 3 shows the performance of each model in terms of its error distributions, which we measured as MAE, MAPE and improvement in MAPE relative to the mechanistic model. All the foundation models performed well on short-term forecasting tasks. Specifically, they slightly outperformed the benchmarks on ILI and influenza and chickenpox, and they substantially outperformed on RSV and dengue. These conclusions are stable across the three different error metrics and also the alternatives presented in the Appendix. Notably, the similar behavior of MAE and MAPE implies that the performance was stable for large and small values. The performance improvement of the foundation models with respect to the benchmarks also seemed more marked for those data, especially dengue, for which available data are scarcer and seasons less regular.

**Figure 3:**
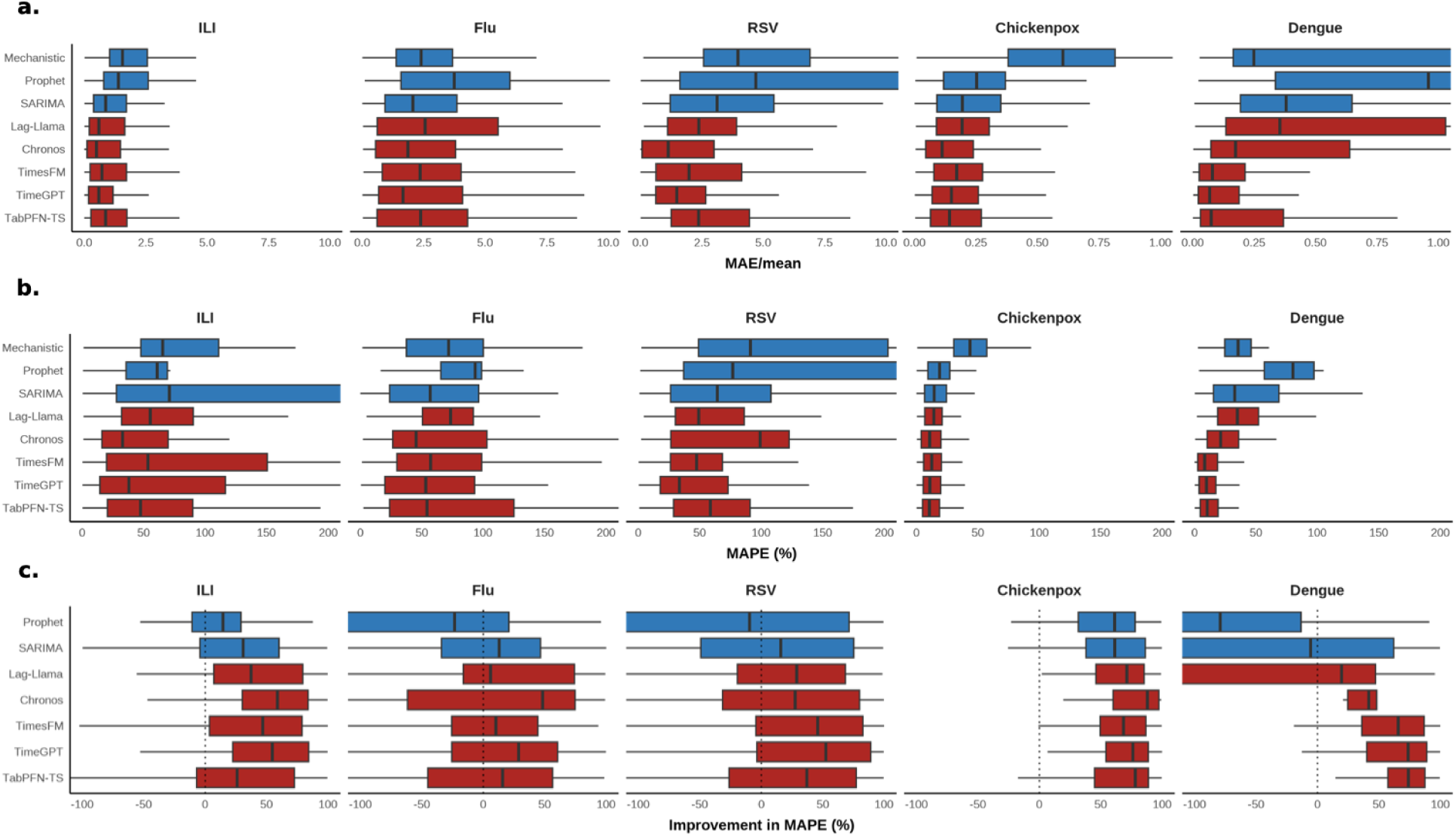
Short-term forecasting. Model performances on short-term (4 week ahead) forecasts, across datasets. Benchmarks are in blue, foundational models are in red. (a) reports MAE divided over the dataset mean. Boxplots indicate the 2.5%, 25%, 50%, 97.5% percentiles. (b) shows the same for MAPE. (c) shows the same for percentage improvement in MAPE.

### Epidemic peak forecasting

We tested the ability of foundation models to forecast the week of occurrence of the seasonal epidemic peak. We considered two scenarios: an early forecast, whereby the forecast was made at the start of the season, and a forecast made 4 weeks before the actual peak. Figure 4a shows the prediction errors of the foundation and the benchmark models in the 2017-2018 seasons. Appendix reports the same for the 2016-2017 and 2018-2019 seasons. Figure 4b compares model performances by ranking them by prediction accuracy.

**Figure 4:**
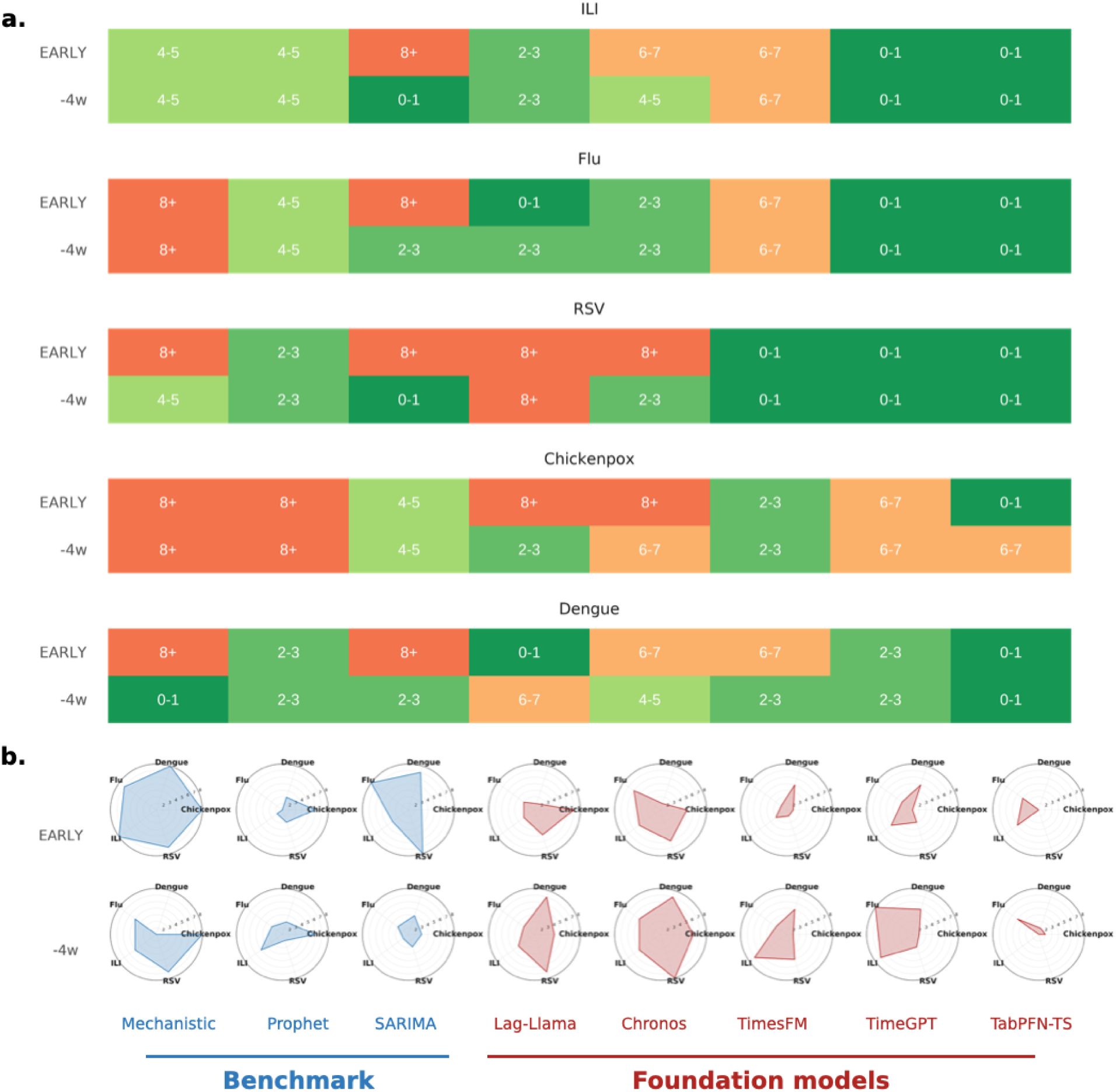
Peak forecasting. **(a)** The plot shows the absolute distance (in weeks) between the estimated peak date and the actual peak date in 2017 for ILI, influenza, RSV, chickenpox, and dengue. The predictions are made from two different time points: EARLY (November of the previous year) and 4 weeks before the real disease peak (-4w). Lower values (green) indicate more accurate peak predictions, while higher values (orange/red) indicate greater deviations. **(b)** Radar chart visualization of median rank of the absolute error in peak timing. The first row represents the ‘EARLY’ forecast horizon, while the second row represents the -4w forecast horizon. Each axis corresponds to a disease, and the rank values compare model performance in predicting the timing of the peak, with lower ranks indicating better performance.

We first review the performance in the short term (4 weeks before the peak). For ILI, TabPFN-TS and TimeGPT tended to perform better than, or equal to the mechanistic model and, generally, any benchmark. The other foundation models instead tended to underperform the mechanistic model. On RSV, Influenza and chickenpox TabPFN-TS, TimeGPT, TimesFM and Lag-Llama had a performance similar to the mechanistic model, while Chronos was slightly worse. For Dengue, foundation models substantially outperformed the mechanistic model in the first two seasons, the opposite in the last season.

The difference in performance between foundation models and benchmark was instead more marked for long-term forecasts (start of the season). TabPFN-TS and TimeGPT consistently outperformed the mechanistic model across all datasets. For Influenza, RSV and chickenpox they substantially outperformed any benchmark, too. For instance, TabPFN-TS forecast the timing of the peak with zero or minimal error (up to one week): this means to accurately forecast the timing of the peak roughly three months in advance. Benchmarks, instead, were mostly unable to give any reliable estimate. Also, Lag-llama forecast the timing of the peak of influenza incidence consistently better than the mechanistic model and most other benchmarks. Foundation models substantially overperformed the benchmark in identifying the timing of the peak of dengue cases, too, across all the seasons.

### Policy evaluation: COVID-19

In late February 2021, COVID-19 incidence in Italy was gradually rising, coinciding with the spread of the Alpha variant - which led to its later dominance - and the continued enforcement of light restrictions (yellow tier) in many regions. This increasing trend ultimately led to the imposition of stricter restrictions (red tier) in several areas. We focused on Latium, the region encompassing Rome, which remained in the yellow tier until March 14, when it was escalated to red. To assess the potential impact of an earlier orange tier adoption, we used TabPFN-TS to estimate the incidence trajectory had the restrictions been tightened earlier - specifically, starting on February 22. Figure 5 presents the incidence predictions generated by TabPFN-TS under two scenarios: the actual tier assignments (top row) and the counterfactual scenario where an orange tier was enforced earlier. The model estimated that this earlier tightening would have averted a median value of 36 detected cases (1st decile: -110, 9th decile: 182) every 100,000 inhabitants in three weeks following its enforcement, to the eventual enforcement of the red tier.

**Figure 5:**
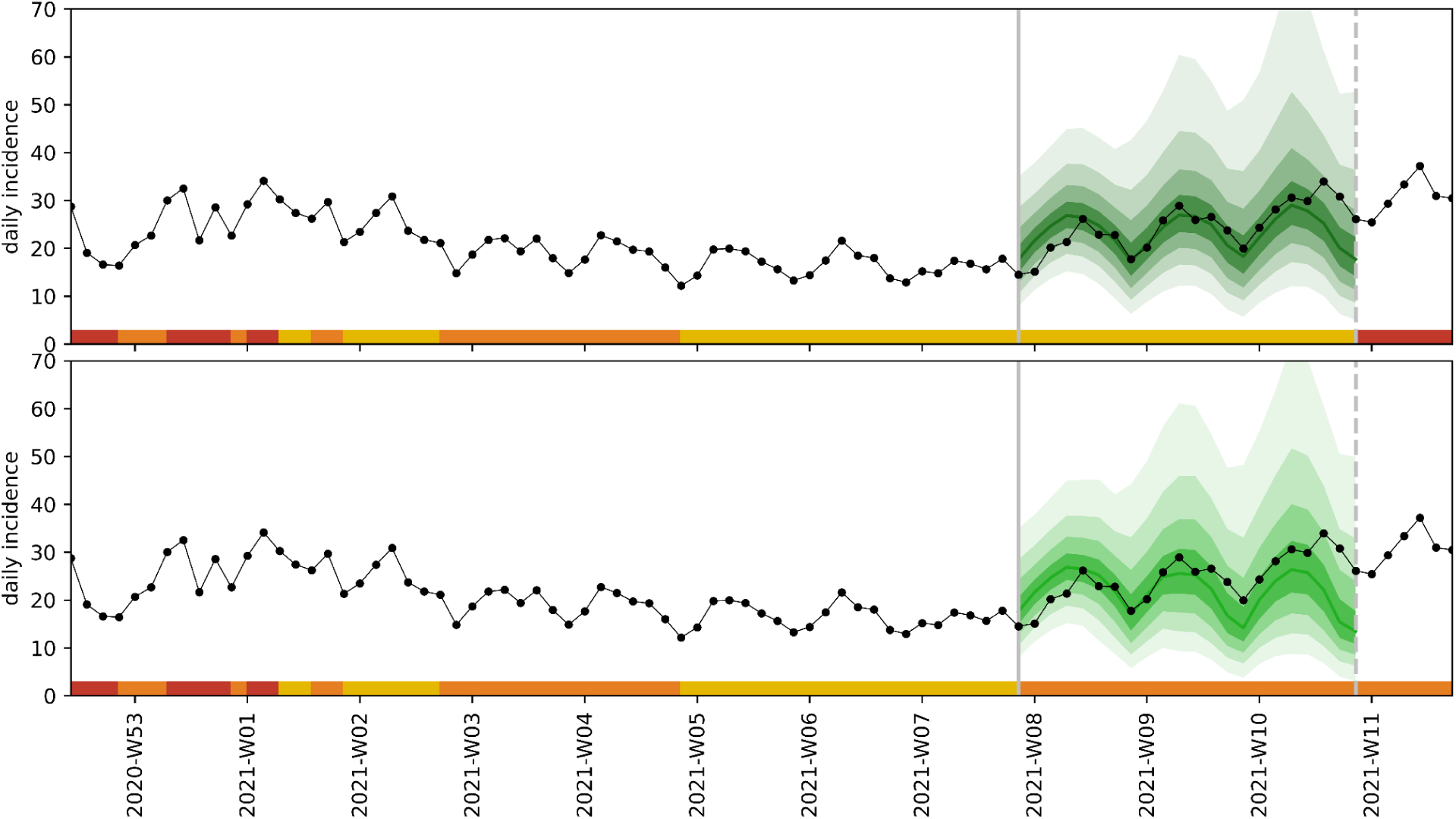
Scenario evaluation: COVID-19 in Latium, Italy. The black line and dots show the daily incidence of cases in Latium, Italy, per 100,000 residents. The bar colored in yellow, orange and red indicates the tier in place. The green curve in the top plot shows the TabPFN-TS prediction assuming the tier enforcement that was actually in place (yellow tier). The green curve in the bottom plot shows the counterfactual prediction assuming that at the start of the window the orange tier would be in place instead of the yellow tier. The colored solid line shows the median prediction, the shaded areas the deciles from 1st to 10th.

### Policy evaluation: neonatal bronchiolitis

The 2023-2024 season marked the introduction in France of newborn immunization against RSV with nirsevimab. We used TabPFN-TS to estimate the impact this immunization campaign had on reducing bronchiolitis-related visits to the ER of infants aged 0-to-12 months. Figure 6 reports the estimated nirsevimab coverage among infants in the general population in three age classes: 0-3 months, 4-6 months and 7-12 months. Coverage changed in these age classes with ongoing immunization and with children getting older. Overall, children aged 0-3 months old had up to 60% immunization from early in the season; the second age class was around 30% for most of the season then increased above 40%. Coverage in the third age class went from around 10% to around 30% along the season. A comparison between the observed ER admission data and the predictions of TabPFN-TS, fine-tuned on four pre-immunization seasons between 2017 and 2023, revealed a significant reduction in admissions in the youngest age class, associated with high nirsevimab coverage. In the second age class the counterfactual without immunization predicted around 50 more ER admissions per fortnight before the season peak, but no difference in peak was measured. The third age class showed the same behavior, albeit with an even lower reduction. Comparison between model predictions and recorded data gave an estimated effectiveness in reducing all-cause bronchiolitis admissions of 56% (1st decile: 10%, 9th decile: 77%), 45% (-20%, 75%) and 13% (-50%, 174%) for the three age classes. While these estimates are not precise due to the study design and the current impossibility to interact with TabPFN-TS’s uncertainty estimates, they are compatible with what was found in a case-control study^42^. Also, notably, TabPFN-TS could correctly estimate the substantially lower effectiveness in the oldest age class found in that study.

**Figure 6:**
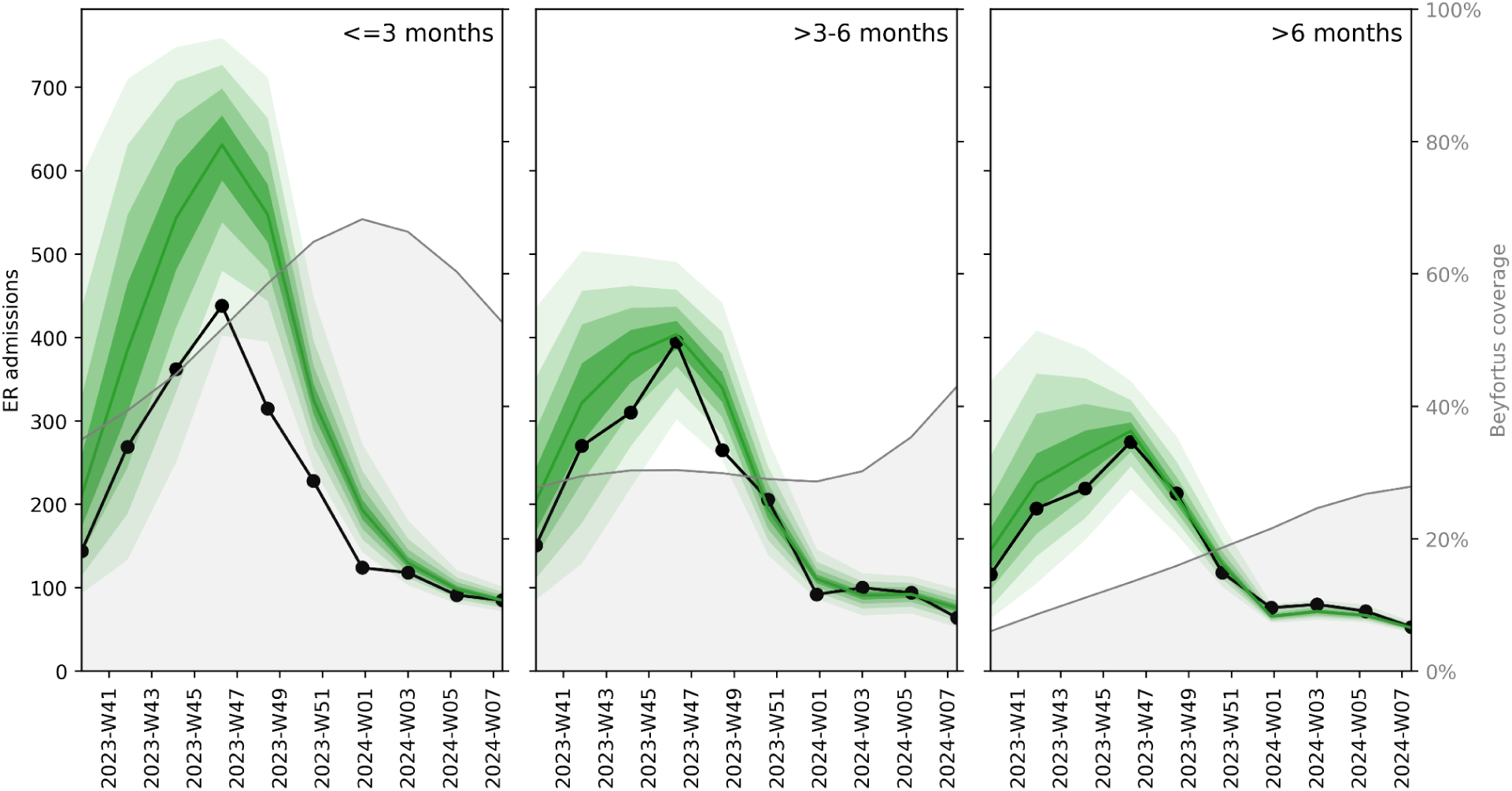
Infant bronchiolitis ER admissions in Paris, France. The black line and dots show the number of fortnightly admissions to the ER due to bronchiolitis in the hospitals under study. The three plots stratify by age of the infant. The green curves show the predictions made by TabPFN-TS in the counterfactual scenario where no RSV immunization was rolled out. The gray lines and filled areas show the expected nirsevimab coverage in that age class.The colored solid line shows the median prediction, the shaded areas the deciles from 1st to 10th.

### Policy evaluation: synthetic scenario

COVID-19 and bronchiolitis demonstrated that TabPFN-TS can generate counterfactual scenarios. The ground truth of these counterfactuals, however, could not be extracted from retrospective data, rendering a validation impossible. For this reason, we tested the performance of TabPFN-TS on policy evaluation on synthetically generated data, too, to assess the accuracy of its prediction in a setting where the counterfactuals could be generated. We used a stochastic SEIRS model with seasonality generating incidence curves in different synthetic regions, with randomly sampled disease and seasonality parameters, to further challenge TabPFN-TS. We modeled randomly enforced interventions as reductions on the transmissibility parameter. Figure S8 showed that TabPFN-TS - fine-tuned on past synthetic data - could correctly forecast incidence of this synthetic epidemic. But most crucially, it showed that it could correctly generate opposite counterfactual scenarios: simulating the effect of interventions that were not enforced, as well as the effect of not enforcing an intervention that was instead enforced, on incidence data. See Appendix for more details.

## Discussion

Our study investigated whether the available foundation AI model for time series can produce accurate forecasts of the evolution of infectious disease epidemics, and inform policies, even when data availability is limited. We tested their performance across different epidemics, diseases and pathogens, transmission routes (airborne, mosquito-borne), locations and indicators (cases from sentinel surveillance, cases from large-scale testing, ER visits).

First, we tested the capability of foundation AI models to generate long-term forecasts, whereby multiple epidemic seasons were predicted. This is a task where traditional modeling approaches typically struggle, unless periodic patterns are very stable and past data are available for training over long periods. Here, this was the case of Influenza-Like-Illness and chickenpox cases in France, where indeed a traditional statistical model like Prophet could extract and reproduce seasonal patterns. But even in these cases, the projected uncertainty of traditional models predictions increased sharply with projection length, rendering multi-season forecasts unreliable. Conversely, the long-term forecasting capabilities of foundation models were striking. Compare the forecast of Prophet and TabPFN-TS on chickenpox data in Figure 2. While both models give accurate median predictions, TabPFN-TS provided substantially lower uncertainty, and, crucially, uncertainty was stable and did not compound over the seasons. Moreover, foundation models provided meaningful forecasts even for epidemics with limited past available data, such as influenza and RSV in France, and dengue in Brazil. In those cases, even when the training data were not enough to completely capture between-season variability, they did forecast crucial elements with reasonable accuracy, such as the time location of the peak.TimeGPT and TimesFM had a performance similar to TabPFN-TS, except TimeGPT failing on RSV. Chronos and Lag-Llama were instead substantially less promising on long-term forecasting: Chronos showed good performance only on chickenpox and could not go beyond one season into the future. Lag-Llama did not perform well across the board.

The performance of the foundation models was also promising on short-term forecasting tasks. There, established models are generally good and widely used for both emergency response and for seasonal epidemic monitoring. But even there, foundation models gave accurate forecasts, possibly outperforming traditional approaches in terms of accuracy and stability, especially in the presence of limited or noisy data.

This ability to generate accurate and stable forecasts with relatively narrow uncertainty bounds, and even when limited data are available, suggests that foundation models could play an essential role in epidemic monitoring and planning both for seasonal epidemic and emergent threats. Given that forecasting consortia typically aggregate multiple models to improve accuracy, we recommend that foundation models start being integrated into ensemble approaches to complement traditional statistical and mechanistic models, to improve overall accuracy, precision and reliability of forecasts^2,9^.

Beyond forecasting epidemic trends, mathematical models are routinely used to evaluate and inform public health policies. Our study showed that foundation models, particularly TabPFN-TS, can generate counterfactual scenarios that can be used to estimate the impact of interventions - or absence thereof - on epidemic indicators such as the incidence of cases or severe disease.

The application to the tiered COVID-19 restrictions in force in Italy in 2021 illustrated this capability: focusing on a period when cases were rising in central Italy concomitantly to the spread of the Alpha variant, but restrictions were minimal, TabPFN-TS could estimate the number of COVID-19 cases that an earlier enforcement of tighter restrictions could have averted. Notably, it could do so with no given knowledge of the pathogen or the disease’s natural history, and training on less than four months of incidence data. This ability to predict the effectiveness of alternative policies in contexts where data are scarce or knowledge of the epidemic are limited - such as the case of an emerging pathogen - is extremely consequential to improve epidemic and pandemic preparedness.

Similarly, our evaluation of the impact of nirsevimab immunization on reducing bronchiolitis-related admissions to the ER demonstrated the potential of foundation models to rapidly evaluate the effectiveness of immunization campaigns. TabPFN-TS could predict an entire season of ER admissions while fine-tuned on very limited data and provide an estimate of age-stratified effectiveness of the immunization campaigns in accordance with an independent epidemiological study^42^. This ability can make TabPFN-TS and other foundation models a precious tool for rapid, near-real-time, policy assessment that can later be corroborated and expanded with larger - but slower, more expensive and logistically more complex - epidemiological studies.

A possible criticism to these conclusions is that validating the counterfactual predictions of these models is challenging, because the ground truth for unimplemented interventions is unknown. To address this, we conducted a validation study using synthetic data generated by a mechanistic epidemic model with known intervention effects. By testing TabPFN-TS on these simulated scenarios, we verified that it could accurately estimate both the direct impact of enforced interventions and the hypothetical impact of interventions that were not implemented.

It should be noted, however, that this kind of validation is often impossible for traditional models, too. Mechanistic models are trusted in the presence of unseen scenarios because of their inherent structure, which reflects the domain knowledge regarding the natural history of the disease and dynamics of the contacts along which it spreads^52^. Foundation models, instead, do not rely on explicit domain knowledge being built in their design, and are not specifically tailored to work on epidemiological data. Nevertheless, further development and extensive testing of the effectiveness of these methods - beyond the scope of this study - will build trust in the scientific community as it was for the established models. In this sense, we are convinced that one major improvement will come through the design of foundation models specifically designed for epidemics forecasting, where domain-specific knowledge will indeed be injected into them - albeit in a new form with respect to mechanistic modeling. This will be possible either by re-training existing foundation models on large-scale epidemic data (both synthetic and real) or by devising new architectures specific to epidemic modeling.

TimeGPT and TabPFN-TS seemed to broadly show the best performance among the foundation models we included. TimeGPT, however, provides only point predictions - no uncertainty - and is not open-source. We therefore identified TabPFN-TS as the most promising among the models tested as it provides accurate, precise and stable predictions, and it is open-source and thus could be developed further. Also, it is extremely lightweight to run and we could fine-tune it on a laptop for all the tasks we tested it on. It should be noted, however, that our study was not comprehensive and we did not test all the available foundation time series models. There are many, and many are being developed^53^. Future studies should investigate whether different model architectures perform better than others for epidemic modeling.

Our study has other limitations. We provided a broad-spectrum performance evaluation of this recently developed modeling approach, showing that it has remarkable potential across epidemics. However, specific studies will be needed to evaluate different foundation models in specific public health settings, before they can be trusted to inform policy making. Also, we included a limited number of benchmarks, while we are aware that many models were developed in the literature for each of the studied epidemic contexts. Our aim, however, was not to prove that foundation time series models are better than any traditional models and should replace them. We showed, instead, that they are promising and they should be the starting point of a new modeling scheme that can integrate existing approaches and improve the overall accuracy and reliability of epidemic modeling as a tool for public health. Notwithstanding, we did show evidence that foundation models are good candidates to outperform traditional models in specific tasks, notably for long-term forecasts and for scenario analysis when limited data are available.

In conclusion, foundation time series models represent a promising new approach for epidemic modeling. Our findings show that these models can produce reliable long- and short-term forecasts, perform well even when data availability is limited, and evaluate the impact of alternative scenarios and the effectiveness of interventions, to inform public health policies. The capacity of these models to generalize across epidemiological contexts, leverage knowledge transfer from non-epidemic data, and operate with minimal computational resources suggests that they could be integrated into routine forecasting efforts and ensemble modeling frameworks. Moreover, their ability to generate counterfactual scenarios with limited data inputs highlights their potential as tools for rapid policy evaluation, particularly in early-stage epidemics and data-scarce settings. As foundation models continue to evolve, their role in public health decision-making will be likely to expand, providing new tools for data-driven epidemic preparedness and response.

## Data Availability

Data on ILI, Chickenpox, Influenza, RSV in France are available from Reseau Sentinelles (https://www.sentiweb.fr/france/fr/?page=database). Data on COVID-19 in Italy are available from https://doi.org/10.1038/s41597-023-02276-y. Bronchiolitis data are from https://doi.org/10.1016/S2352-4642(24)00171-8.

## Data sharing

Data on ILI, Chickenpox, Influenza, RSV in France are available from Réseau Sentinelles (https://www.sentiweb.fr/france/fr/?page=database). Data on COVID-19 in Italy are available from Ref.^40^.

## Acknowledgements

This study was partially supported by Horizon Europe grant SIESTA (101131957) to E.V., the CIPHOD project (ANR-23-CPJ1-0212-01) to F. B., the European Union’s Horizon Europe research and innovation programme Cofund SOUND.AI under the Marie Sklodowska-Curie Grant Agreement No 101081674 to S.K.

## Notes

### Competing Interest Statement

The authors have declared no competing interest.

### Author Declarations

No individual data were used here, not even in an anonymized form. Only population-level case count were used. Data on ILI, Chickenpox, Influenza, RSV in France are available from Reseau Sentinelles (https://www.sentiweb.fr/france/fr/?page=database). Data on COVID-19 in Italy are available from https://doi.org/10.1038/s41597-023-02276-y. Bronchiolitis data are from https://doi.org/10.1016/S2352-4642(24)00171-8.

