## Appendix for "Foundation time series models for forecasting and policy evaluation in infectious disease epidemics"

Suprabhath Kalahasti[1], Benjamin Faucher[1], Boxuan Wang[1], Claudio Ascione[1], Ricardo Carbajal[2], Maxime Enault[2], Christophe Vincent Cassis[2], Titouan Launay[1], Caroline Guerrisi[1], Pierre-Yves Boëlle[1], Federico Baldo[1], Eugenio Valdano[1]*

[1]Sorbonne Université, INSERM, Institut Pierre Louis d'Epidémiologie et de Santé Publique, F75012, Paris, France.
[2] AP-HP, Hôpital Armand Trousseau, Paediatric Emergency Department, Paris, France.

**Supplementary Text** **2**
- Benchmark: Mechanistic model 2
- Evaluation score 4
- Scenarios for long-term, short-term forecasting and peak estimation. 5
- Policy evaluation: synthetic scenario 7
  - Generation 7
  - Inference 8

**Supplementary Figures** **9**
- Short-term forecasting 9
- Peak forecasting 11
- Policy evaluation 13
- References 15

### Supplementary Text

#### Benchmark: Mechanistic model

We used a modified Susceptible-Infected-Recovered (SIR) model with seasonal dynamics to fit the data and realize forecasts. The model is described by the following equations.

$$\frac{dS}{dt} = -\beta(t).S(t).I(t)/N$$

$$\frac{dI}{dt} = \beta(t).S(t).I(t)/N - \mu.I(t)$$

$$\frac{dR}{dt} = \mu.I(t)$$

With $\beta(t) = \beta_0.[1 + \delta.cos(\omega t + \phi)]$

The parameters are summarized in Tab. S1.

| | | |
|---|---|---|
| $\beta_0$ | Baseline transmission rate | To be estimated |
| $\mu$ | Recovery rate | ILI : 1/6[1]<br>Flu : 1/6[1]<br>RSV : 1/8[2]<br>Dengue : 1/10[3]<br>Chickenpox : 1/7[4] |
| $R(t = 0)$ | Initial number of recovered | To be estimated |
| $I(t = 0)$ | Initial number of infected | To be estimated |
| $\omega$ | Period of seasonality | $2\pi/365$ |
| $\phi$ | Phase of seasonality | To be estimated |
| $\delta$ | Force of seasonality | To be estimated |

**Table S1** Parameters of the SIR model.

For a disease D, for the $N_D$ season prior to 2016, we fitted the complete set of parameters $\Theta_D = \{\beta_k, \phi_k, \delta_k, R(t=0)_k, I(t=0)_k \mid k \in [[1, N_D]]\}$, over the entire season. The fit of seasons until 2016 for ILI is shown as an example in Fig. S1. This approach yielded to $N_D$ estimates of each parameter, from which we computed the mean and standard deviation. We then applied the calibrated model to the 2016–2017 season, fitting the SIR model to the early-season data up to a time point $t_0$, depending on the disease and the scenario (see Tab. S1).

The model fitting was performed using Variational Inference for Approximated Complex Bayesian Posterior Distribution with the CmdStanPy package. This method allows for a computationally efficient estimation of posterior distributions while capturing correlations between parameters. The variational inference procedure was initialized using the mean parameter values computed from past seasons on $\Theta_D$. Priors were specified as normal distributions, with means and standard deviations computed on $\Theta_D$. This approach ensured that the inference was informed by historical patterns while allowing flexibility for seasonal variations.

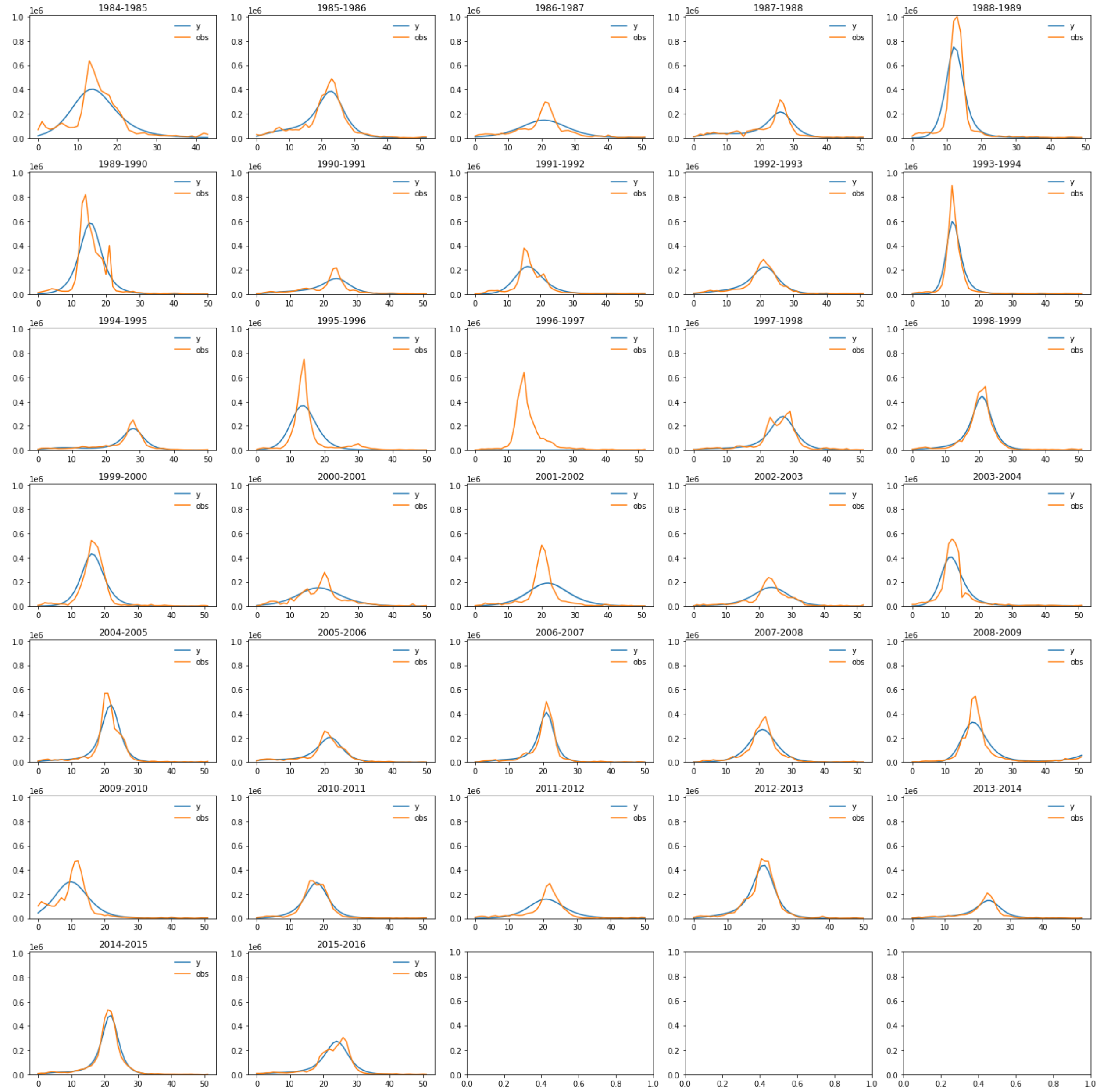


**Figure S1** Fit of seasons before 2016 (from 1984–1985 to 2015–2016) with the mechanistic model. Orange lines represent the data, while blue lines represent the incidence estimated by the model.

#### Evaluation score

We evaluate the models using metrics that measure both point forecast accuracy and relative performance. Different metrics are applied based on the type of forecasting task.

**Mean Absolute Error (MAE):** Given a forecast $\hat{y}_i$ and an actual corresponding value $y_i$, the mean absolute error is computed as:

$$\text{MAE} = \frac{1}{n}\sum_{i=1}^{n}|y_i - \hat{y}_i|$$

This metric measures the average magnitude of forecast errors without considering their direction. It provides a straightforward measure of how close the forecasts are to the observed values.

**Mean Absolute Percentage Error (MAPE):** MAPE measures the percentage difference between the forecast and the actual value, making it useful for comparisons across diseases of different incidence scales:

$$\text{MAPE} = \frac{100\%}{n}\sum_{i=1}^{n}\left|\frac{y_i - \hat{y}_i}{y_i}\right|$$

**Absolute Error(AE):** AE is used for evaluating models that predict the peak timing and magnitude. It provides a straightforward measure of the deviation between predicted and actual values.

Peak Timing Error: Given a predicted peak date $t_{\text{predicted}}$ and the actual peak date $t_{\text{actual}}$, the absolute error in timing is computed as:

$$\text{Timing Error} = |t_{\text{predicted}} - t_{\text{actual}}|$$

#### Scenarios for long-term, short-term forecasting and peak estimation.

We summarize in Tab. S2 the starting points of the forecast for scenarios explored in Fig 2, 3 and 4 of the main paper depending on the disease.

| Scenario | ILI | Flu | RSV | Chickenpox | Dengue |
|---|---|---|---|---|---|
| **Long term forecast A** | 2016-10-03 | 2016-10-03 | 2016-10-03 | 2016-09-05 | 2016-10-03 |
| **Long term forecast B** | 2016-11-07 | 2016-11-07 | 2016-11-07 | 2016-10-03 | 2016-11-07 |
| **4 week forecast from peak 2016 minus 4 weeks** | 2016-12-19 | 2016-12-19 | 2016-11-28 | 2017-02-20 | 2017-02-19 |
| **4 week forecast from peak 2016 minus 3 weeks** | 2016-12-26 | 2016-12-26 | 2016-12-05 | 2017-02-27 | 2017-02-26 |
| **4 week forecast from peak 2016 minus 2 weeks** | 2017-01-02 | 2017-01-02 | 2016-12-12 | 2017-03-06 | 2017-03-05 |
| **4 week forecast from peak 2016 minus 1 weeks** | 2017-01-09 | 2017-01-09 | 2016-12-19 | 2017-03-13 | 2017-03-12 |
| **4 week forecast from peak 2016** | 2017-01-16 | 2017-01-16 | 2016-12-26 | 2017-03-20 | 2017-03-19 |
| **4 week forecast from peak 2016 plus 1 weeks** | 2017-01-23 | 2017-01-23 | 2017-01-02 | 2017-03-27 | 2017-03-26 |

| | | | | | |
|---|---|---|---|---|---|
| **4 week forecast from peak 2016 plus 2 weeks** | 2017-01-30 | 2017-01-30 | 2017-01-09 | 2017-04-03 | 2017-04-02 |
| **4 week forecast peak 2016 plus 3 weeks** | 2017-02-06 | 2017-02-06 | 2017-01-16 | 2017-04-10 | 2017-04-09 |
| **4 week forecast from peak 2017 minus 4 weeks** | 2017-02-13 | 2017-02-13 | 2017-01-23 | 2017-04-17 | 2017-04-16 |
| **4 week forecast from peak 2017 minus 4 weeks** | 2017-11-27 | 2017-11-27 | 2017-11-27 | 2018-02-19 | 2018-03-11 |
| **4 week forecast from peak 2017 minus 3 weeks** | 2017-12-04 | 2017-12-04 | 2017-12-04 | 2018-02-26 | 2018-03-18 |
| **4 week forecast from peak 2017 minus 2 weeks** | 2017-12-11 | 2017-12-11 | 2017-12-11 | 2018-03-05 | 2018-03-25 |
| **4 week forecast from peak 2017 minus 1 weeks** | 2017-12-18 | 2017-12-18 | 2017-12-18 | 2018-03-12 | 2018-04-01 |
| **4 week forecast from peak 2017** | 2017-12-25 | 2017-12-25 | 2017-12-25 | 2018-03-19 | 2018-04-08 |
| **4 week forecast from peak 2017 plus 1 weeks** | 2018-01-01 | 2018-01-01 | 2018-01-01 | 2018-03-26 | 2018-04-15 |
| **4 week forecast from peak 2017 plus 2 weeks** | 2018-01-08 | 2018-01-08 | 2018-01-08 | 2018-04-02 | 2018-04-22 |
| **4 week forecast peak 2017 plus 3 weeks** | 2018-01-15 | 2018-01-15 | 2018-01-15 | 2018-04-09 | 2018-04-29 |
| **4 week forecast peak 2017 plus 4 weeks** | 2018-01-22 | 2018-01-22 | 2018-01-22 | 2018-04-16 | 2018-05-06 |
| **4 week forecast from peak 2018 minus 4 weeks** | 2019-01-14 | 2018-12-31 | 2018-12-31 | 2019-03-11 | 2019-03-17 |
| **4 week forecast from peak 2018 minus 3 weeks** | 2019-01-21 | 2019-01-07 | 2019-01-07 | 2019-03-18 | 2019-03-24 |
| **4 week forecast from peak 2018 minus 2 weeks** | 2019-01-28 | 2019-01-14 | 2019-01-14 | 2019-03-25 | 2019-03-31 |
| **4 week forecast from peak 2018 minus 1 weeks** | 2019-02-04 | 2019-01-21 | 2019-01-21 | 2019-04-01 | 2019-04-07 |
| **4 week forecast from peak 2018** | 2019-02-11 | 2019-01-28 | 2019-01-28 | 2019-04-08 | 2019-04-14 |
| **4 week forecast from peak 2018 plus 1 weeks** | 2019-02-18 | 2019-02-04 | 2019-02-04 | 2019-04-15 | 2019-04-21 |
| **4 week forecast from peak 2018 plus 2 weeks** | 2019-02-25 | 2019-02-11 | 2019-02-11 | 2019-04-22 | 2019-04-28 |
| **4 week forecast peak 2018 plus 3 weeks** | 2019-03-04 | 2019-02-18 | 2019-02-18 | 2019-04-29 | 2019-05-05 |
| **4 week forecast peak 2018 plus 4 weeks** | 2019-03-11 | 2019-02-25 | 2019-02-25 | 2019-05-06 | 2019-05-12 |
| **Peak estimation 2016 A** | 2016-10-03 | 2016-10-03 | 2016-10-03 | 2016-09-05 | 2016-10-03 |
| **Peak estimation 2016 B** | 2016-11-07 | 2016-11-07 | 2016-11-07 | 2016-10-03 | 2016-11-07 |
| **Peak estimation 2017 A** | 2017-10-02 | 2017-10-02 | 2017-10-02 | 2017-07-03 | 2017-10-02 |
| **Peak estimation 2017 B** | 2017-11-06 | 2017-11-06 | 2017-11-06 | 2017-08-07 | 2017-11-06 |
| **Peak estimation 2018 A** | 2018-10-01 | 2018-10-01 | 2018-10-01 | 2018-07-02 | 2018-10-01 |

| | | | | | |
|---|---|---|---|---|---|
| **Peak estimation 2018 B** | 2018-11-05 | 2018-11-05 | 2018-11-05 | 2018-08-06 | 2018-11-05 |
| **Peak estimation 2016 from 4 weeks before peak** | 2016-10-03 | 2016-12-19 | 2016-11-28 | 2017-02-20 | 2017-02-19 |
| **Peak estimation 2016 from 3 weeks before peak** | 2016-11-07 | 2016-12-26 | 2016-12-05 | 2017-02-27 | 2017-02-26 |
| **Peak estimation 2016 from 2 weeks before peak** | 2016-10-03 | 2017-01-02 | 2016-12-12 | 2017-03-06 | 2017-03-05 |
| **Peak estimation 2016 from 1 week before peak** | 2016-11-07 | 2017-01-09 | 2016-12-19 | 2017-03-13 | 2017-03-12 |
| **Peak estimation 2017 from 4 weeks before peak** | 2016-10-03 | 2017-11-27 | 2017-11-27 | 2018-02-19 | 2018-03-11 |
| **Peak estimation 2017 from 3 weeks before peak** | 2016-11-07 | 2017-12-04 | 2017-12-04 | 2018-02-26 | 2018-03-18 |
| **Peak estimation 2017 from 2 weeks before peak** | 2016-10-03 | 2017-12-11 | 2017-12-11 | 2018-03-05 | 2018-03-25 |
| **Peak estimation 2017 from 1 week before peak** | 2016-11-07 | 2017-12-18 | 2017-12-18 | 2018-03-12 | 2018-04-01 |
| **Peak estimation 2018 from 4 weeks before peak** | 2016-10-03 | 2018-12-31 | 2018-12-31 | 2019-03-11 | 2019-03-17 |
| **Peak estimation 2018 from 3 weeks before peak** | 2016-11-07 | 2019-01-07 | 2019-01-07 | 2019-03-18 | 2019-03-24 |
| **Peak estimation 2018 from 2 weeks before peak** | 2016-10-03 | 2019-01-14 | 2019-01-14 | 2019-03-25 | 2019-03-31 |
| **Peak estimation 2018 from 1 week before peak** | 2016-11-07 | 2019-01-21 | 2019-01-21 | 2019-04-01 | 2019-04-07 |

**Table S2** Forecast starting points based on the scenario and disease.

#### Policy evaluation: synthetic scenario

##### Generation

We implemented a stochastic, discrete-time SEIRS model to simulate the transmission dynamics of an infectious disease in a single population, incorporating seasonality and interventions. The model tracks four compartments: Susceptible (S), Exposed (E), Infected (I), and Recovered (R), with transitions governed by binomially distributed probabilities. We assume a constant population of 100,000 individuals. The model features a transmission rate $\beta$ that tunes the force of infection as customary: $\beta I/N$. It also features a E-to-I rate $\epsilon$ tuning the latency period, a recovery rate $\mu$ and a waning immunity rate $\omega$. The model parameters were set as follows: $\epsilon = 0.27$ corresponding to an average latency period of 3.7 days[1] and $\omega = 0.01$ corresponding to an average duration of immunity of 3 months. Seasonality was introduced through a periodic modulation of the transmission rate, modeled as $\beta(t) = \beta_0(1 + A\sin(2\pi t/T)$, with T=365 days. We generated epidemics in 5 synthetic regions, in each sampling $\beta_0$ uniformly in [0.52, 0.56] and $\mu$ in [0.26, 0.28], corresponding to an average infectious period of roughly 3.7 days. Seasonal forcing A was sampled uniformly in [0.15, 0.2]. Interventions were modeled as time-dependent modifications to $\beta(t)$, represented by step functions that activate and deactivate at predefined times. Interventions were implemented at random times and lasted a random duration between 1 and 3 months. They entailed a reduction on transmission randomly sampled in [-2%, -20%]. For each of these synthetic regions we generated 10 years of data. We remark that no information on disease parameters, seasonality and the strength of interventions was given to TabPFM, but only when interventions were in place.

#### Inference

We used TabPFM to generate counterfactual scenarios to study the impact of an intervention on the incidence of infections. Specifically, we simulated the enforcement of an intervention that was not present when the data were generated, and its opposite: the absence of an intervention that was enforced at data generation. We focused on one synthetic region and, for each prediction task we fine-tuned TabPFM using data from all the synthetic regions up to the last available date prior to the prediction, similar to what we did for COVID-19 in Italian regions. The enforcement of interventions was modeled through a binary feature being equal to zero for days without interventions, and one for days with interventions. During weekly data aggregation, the intervention variable was averaged over the week. Figure S8 shows that TabPFM can correctly estimate the number of infections that an intervention averted. This is visible by noticing that the blue curve correctly forecasts the effect of the interventions around week 36 of year 8 and week 18 of year 9, while the red curve predicts more infections that what was observed. At the same time, TabPFM can also estimate the impact of estimating the reduction in incidence between week 53 of year 9 and week 9 of year 10 that would have occurred (green scenario) if an intervention had been in place. Instead, the orange scenario, where the intervention is not present, correctly tracks observed infections.

### Supplementary Figures

#### Short-term forecasting

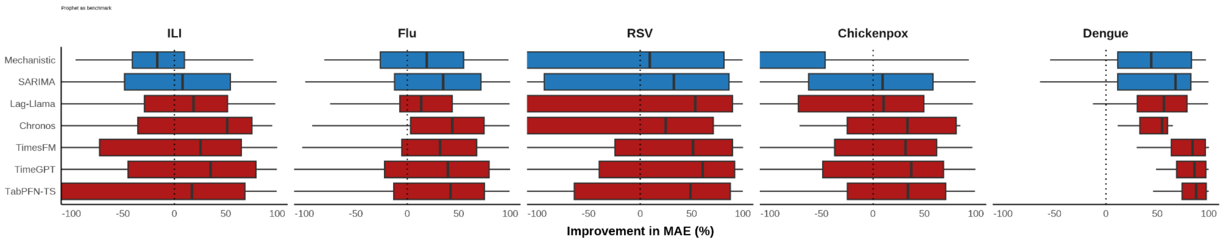


**Figure S2: MAE Improvement of Forecasting Models Relative to Prophet.** This figure illustrates the percentage improvement (or decline) in the Mean Absolute Error (MAE) of various forecasting models compared to the Prophet model across five diseases: ILI, Flu, RSV, Chickenpox, and Dengue. Benchmark models (blue) include Mechanistic and SARIMA, while foundation models (red) include LLM-based models, Chronos, TimeGPT, and TabPFN-TS. Positive values indicate improved forecasting performance, while negative values suggest a decline relative to Prophet.

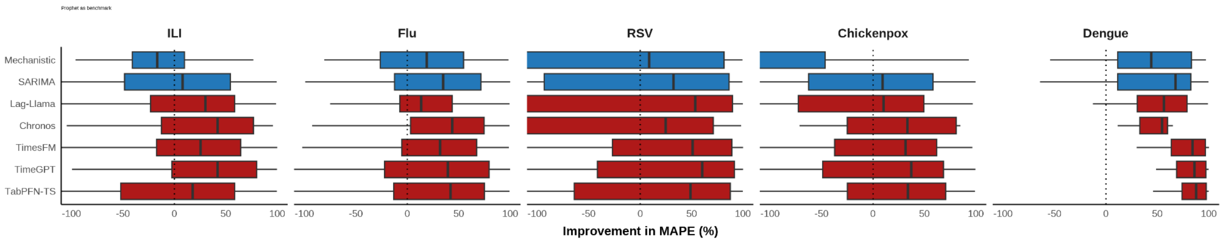


**Figure S3: MAPE Improvement of Forecasting Models Relative to Prophet.** This figure illustrates the percentage improvement (or decline) in Mean Absolute Percentage Error (MAPE) of various forecasting models compared to the Prophet model across five diseases: ILI, Flu, RSV, Chickenpox, and Dengue. Benchmark models (blue) include Mechanistic and SARIMA, while foundation models (red) include LLM-based models, Chronos, TimeGPT, and TabPFN-TS. Positive values indicate improved forecasting performance, while negative values suggest a decline relative to Prophet.

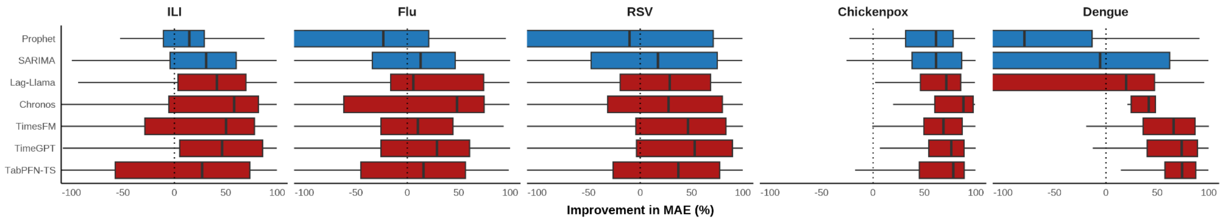

**Figure S4: MAPE Improvement of Forecasting Models Relative to Mechanistic model.** This figure illustrates the percentage improvement (or decline) in Mean Absolute Percentage Error (MAPE) of various forecasting models compared to the Mechanistic model across five diseases: ILI, Flu, RSV, Chickenpox, and Dengue. Benchmark models (blue) include Prophet and SARIMA, while foundation models (red) include LLM-based models, Chronos, TimeGPT, and TabPFN-TS. Positive values indicate improved forecasting performance, while negative values suggest a decline relative to Prophet.

#### Peak forecasting

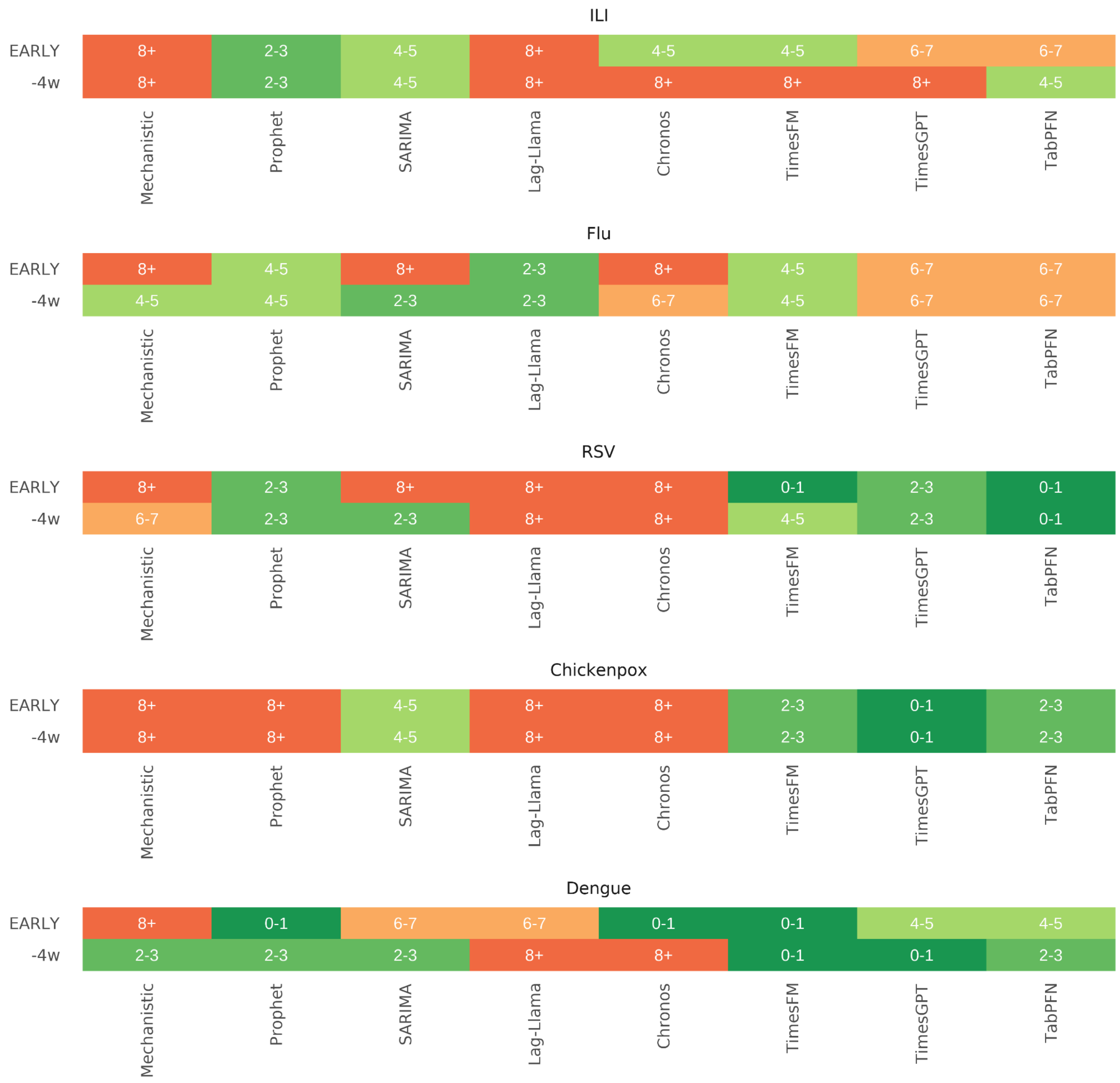


**Figure S5: Absolute Distance Between Predicted and Actual Peak Dates for Different Diseases (2016).** This heatmap shows the absolute distance (in weeks) between the estimated peak date and the actual peak date for five diseases: ILI, Flu, RSV, Chickenpox, and Dengue. The predictions are made from two different time points: EARLY (November of the previous year) and 4 weeks before the real disease peak. Lower values (green) indicate more accurate peak predictions, while higher values (orange/red) indicate greater deviations. The results illustrate how prediction accuracy varies across diseases and forecasting timeframes.

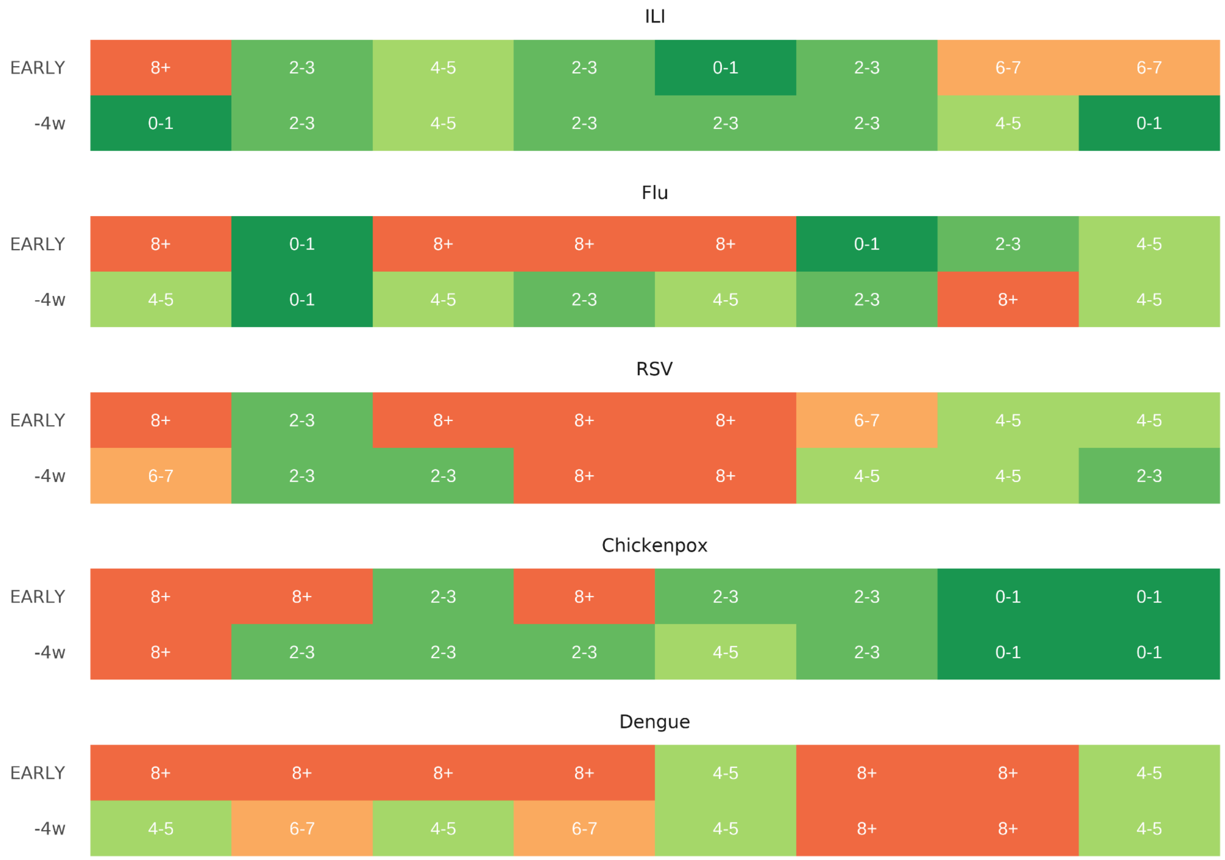


**Figure S6: Absolute Distance Between Predicted and Actual Peak Dates for Different Diseases (2018).** This heatmap shows the absolute distance (in weeks) between the estimated peak date and the actual peak date for five diseases: ILI, Flu, RSV, Chickenpox, and Dengue. The predictions are made from two different time points: EARLY (November of the previous year) and 4 weeks before the real disease peak. Lower values (green) indicate more accurate peak predictions, while higher values (orange/red) indicate greater deviations. The results illustrate how prediction accuracy varies across diseases and forecasting timeframes.

#### Policy evaluation

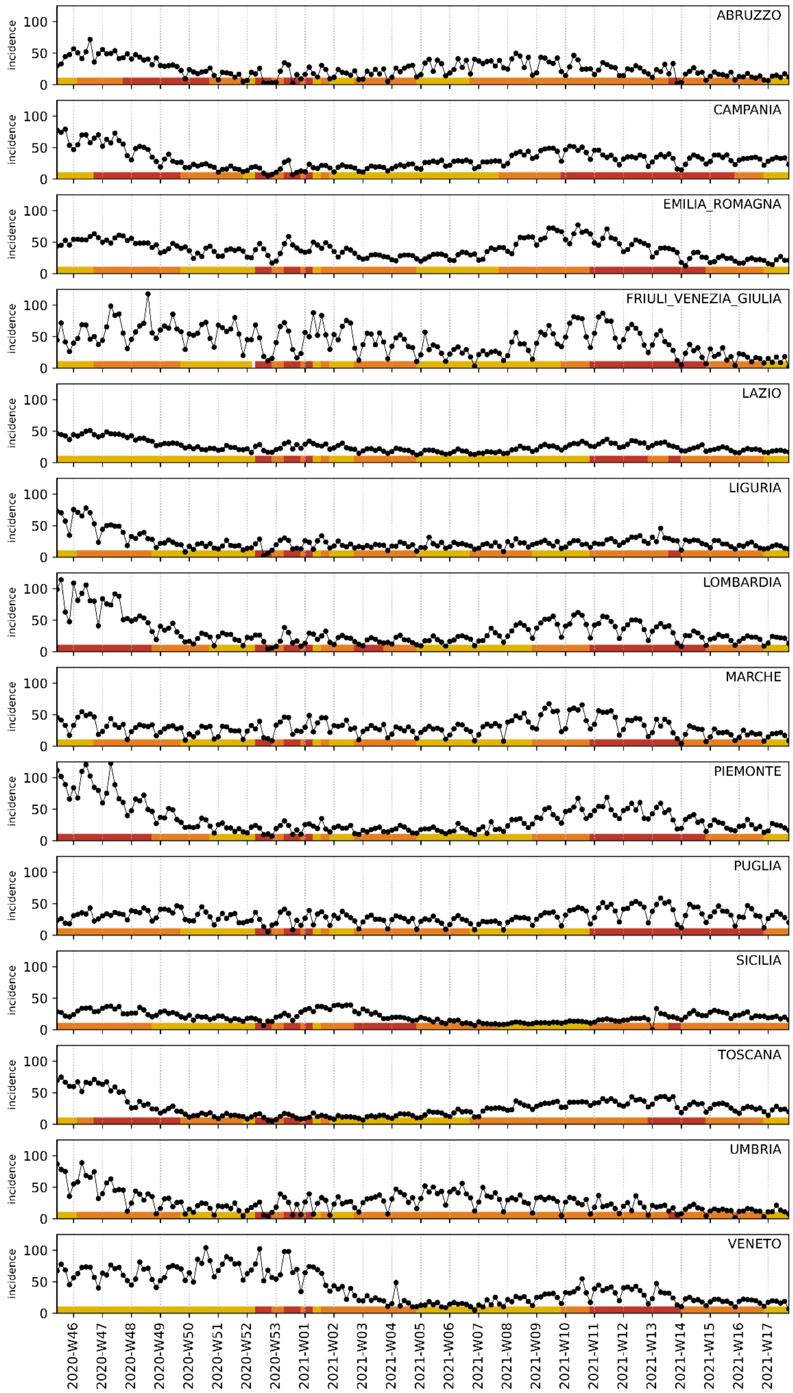


**Figure S7: Daily incidence of cases of COVID-19 in selected Italian regions.** Data are from November 2020 to May 2021. Incidence is the number of detected cases per 100,000 residents. The imposed restriction tier is indicated with the color (yellow, orange, red).

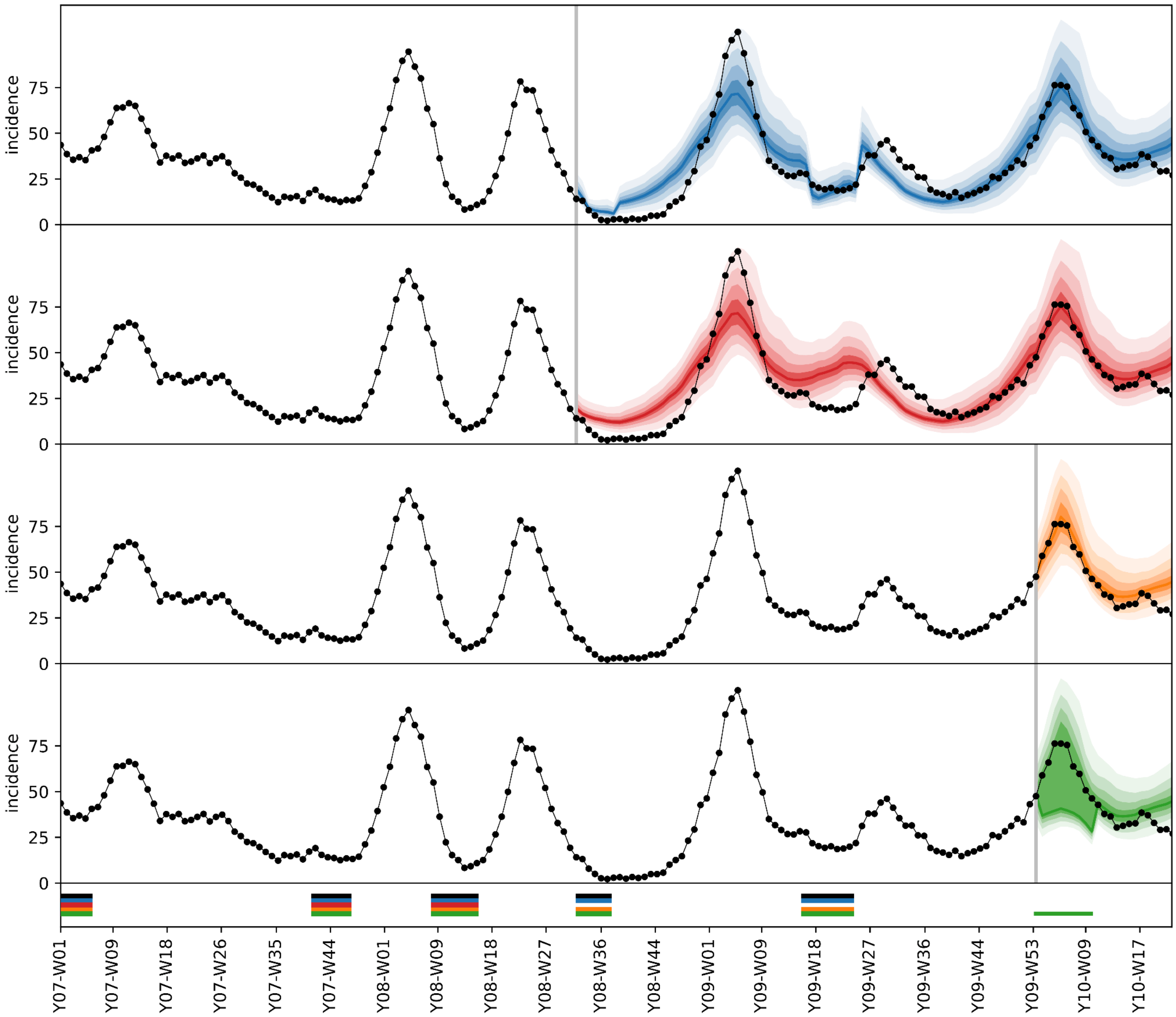


**Figure S8: Policy evaluation - synthetic scenario.** The four large panels report the same incidence data (black dots and curve) generated in one synthetic country, and four different prediction scenarios. Gray vertical bars indicate the start of prediction windows. The bottom narrow panel indicates what interventions were present at data generation (black bar) and whether they were fed to each of the scenarios. The blue and orange scenarios include all interventions that were used to generate the data, and only those. The red scenario includes no interventions after the prediction window, even when they were used at data generation.The green scenario includes one late intervention which was not included at data generation. Predictions show all the deciles from the 1st to the 9th. The solid central line represents the median (5th decile).
